# Experimental human pneumococcal colonisation in older adults is feasible and safe, not immunogenic

**DOI:** 10.1101/2020.04.23.20077073

**Authors:** Hugh Adler, Esther L German, Elena Mitsi, Elissavet Nikolaou, Sherin Pojar, Caz Hales, Rachel Robinson, Victoria Connor, Helen Hill, Angela D Hyder-Wright, Lepa Lazarova, Catherine Lowe, Emma L Smith, India Wheeler, Seher R Zaidi, Simon P Jochems, Dessi Loukov, Jesús Reiné, Carla Solórzano-Gonzalez, Polly de Gorguette d’Argoeuves, Tessa Jones, David Goldblatt, Tao Chen, Stephen J Aston, Neil French, Andrea M Collins, Stephen B Gordon, Daniela M Ferreira, Jamie Rylance

**Author notes:** Corresponding authors: Dr Jamie Rylance, Prof Daniela Ferreira, (Respiratory Infection Group, Liverpool School of Tropical Medicine, Liverpool L3 5QA, UK). These authors are joint last authors (Daniela M Ferreira, Jamie Rylance). Despite low rates of pneumococcal colonisation detected in older people in previous community surveys, experimental colonisation was established in 39% of volunteers aged ≥ 50 years. The serological and functional immune responses of older adults to challenge and colonisation with live pneumococci were markedly different to those of young adults. Experimental human pneumococcal colonisation was safe, supporting the use of this methodology in clinical trials of pneumococcal vaccines for older people. ***Declarations:*** This study was approved by the National Health Service Research Ethics Committee (Ref 16/NW/0031), registered with ISRCTN (10948363) and funded by the Bill and Melinda Gates Foundation (Grant number OPP1117728) and UK Medical Research Council (Grant number MR/M011569/1). The funders had no role in study design, data analysis or the decision to submit for publication. This article has an online data supplement, which is accessible from this issue’s table of content online at www.atsjournals.org. **Contributions**. **Obtained funding:** SBG, DMF. **Study design:** HA, SBG, DMF, J Rylance. **Obtained ethical approval:** HA, J Rylance. **Participant recruitment, clinical procedures and data collection:** HA, CH, RR, VC, HH, ADH-W, LL, CL, ES, IW, SRZ. **Data analyses:** HA, SP, ELG, SPJ, EM, EN, DL, JR, CSG, PGA, TJ, DG, TC, DF. **Study oversight:** AMC, SJA, NF, SBG, DMF, J Rylance. All authors contributed to the manuscript and approved the final version submitted for publication.

## Abstract

**Rationale:** Pneumococcal colonisation is key to the pathogenesis of invasive disease, but is also immunogenic in young adults, protecting against re-colonisation. Colonisation is rarely detected in older adults, despite high rates of pneumococcal disease.

**Objectives:** To establish experimental human pneumococcal colonisation in healthy adults aged 50—84 years, to measure the immune response to pneumococcal challenge, and to assess the protective effect of prior colonisation against autologous strain rechallenge.

**Methods:** Sixty-four participants were inoculated with *Streptococcus pneumoniae* (serotype 6B, 80,000CFU in each nostril). Colonisation was determined by bacterial culture of nasal wash, serum anti-6B capsular IgG responses by ELISA, and anti-protein immune responses by multiplex electrochemiluminescence.

**Measurements and Main Results:** Experimental colonisation was established in 39% of participants (25/64) with no adverse events. Colonisation occurred in 47% (9/19) of participants aged 50—59 compared with 21% (3/14) in those aged ≥70 years. Previous pneumococcal polysaccharide vaccination did not protect against colonisation. Colonisation did not confer serotype-specific immune boosting: GMT (95% CI) 2.7μg/mL (1.9—3.8) pre-challenge versus 3.0 (1.9—4.7) four weeks post-colonisation (p = 0.53). Furthermore, pneumococcal challenge without colonisation led to a drop in specific antibody levels from 2.8μg/mL (2.0—3.9) to 2.2μg/mL (1.6—3.0) post-challenge (p = 0.006). Anti-protein antibody levels increased following successful colonisation. Rechallenge with the same strain after a median of 8.5 months (IQR 6.7—10.1) led to recolonisation in 5/16 (31%).

**Conclusions:** In older adults, experimental pneumococcal colonisation is feasible and safe, but demonstrates different immunological outcomes compared with younger adults in previous studies.

## Introduction

Older adults have high rates of morbidity and mortality from pneumococcal disease, including pneumonia and meningitis (1-4). Nasopharyngeal pneumococcal colonisation necessarily precedes pneumococcal disease (5). Paradoxically, colonisation is infrequently detected in older adults, with rates ranging from 1.9% to 4.2% in studies employing culture-based methodology (6-8). This may be due to insensitive detection methods or an altered niche of colonisation in older adults (9, 10). Others have suggested that older adults are inherently less susceptible to colonisation, perhaps due to age-related dysregulation of airway inflammatory pathways (11-13). Colonisation is immunogenic in young adults, protecting against future recolonisation with the same serotype (14, 15), and therefore reduced colonisation in older adults may partially explain their susceptibility to pneumococcal disease. The immune dynamics of colonisation in older adults have not been previously studied.

In adults over 65 years of age, recommendations for pneumococcal polysaccharide vaccine (PPV23) and 13-valent pneumococcal conjugate vaccine (PCV13) administration vary between countries (16, 17). Both vaccines induce anti-capsular polysaccharide antibody production, although the functionality of antibodies produced is reduced in older adults (18, 19). Both vaccines confer a degree of protection against vaccine-serotype invasive pneumococcal disease (20, 21), although a combination of imperfect efficacy and serotype replacement by non-vaccine serotypes have reduced the effectiveness of both vaccines (22). PCV13 confers short-term protection against colonisation (23), most likely via antibody-mediated bacterial agglutination (24), while high-quality prospective data regarding PPV23 and colonisation are lacking (25). Protection against colonisation is a surrogate for protection against disease. Since neither vaccine confers complete protection against pneumococcal disease in older adults, improved vaccines and immunisation strategies are urgently required.

Controlled human infection models have revolutionised the study of infectious disease pathogenesis (26). Such studies typically recruit healthy, young adults who are at low risk of severe disease or complications. Rhinovirus challenge studies have been performed in older participants, including those with respiratory comorbidities (27), but bacterial challenge studies have not been systematically attempted in older people. There is an unmet need for such research in this population, given the higher age-specific incidence of pathogens such as pneumococcus, and the uncertain generalisability from studies of young adults.

We report the expansion of the experimental human pneumococcal colonisation (EHPC) model into adults aged ≥50 years, aiming to establish the feasibility, susceptibility and safety of EHPC in older adults. We also define the immunogenicity of pneumococcal colonisation in this population measured by antibody responses, and the protection conferred by primary challenge against subsequent re-colonisation by the same strain.

## Methods

Healthy adults aged 50—84 years were approached using advertisements, research volunteer databases and primary care patient lists. All participants provided written informed consent and underwent safety screening including review of primary care records, physical examination, electrocardiography, spirometry, complete blood count and biochemical profile before participation (see Supplementary Appendix for full inclusion/exclusion criteria and study methods). In brief, participants were excluded if they had regular close contact with children aged <5 years or immunosuppressed adults, uncontrolled medical comorbidity, recent steroid or antibiotic therapy, significant smoking history, or history of culture-proven pneumococcal disease. Vaccination history was recorded: in the UK, PPV23 is recommended to all 65-year olds, but PCV13 is not routinely offered. The study was overseen by an independent Data Monitoring and Safety Committee. Experimental pneumococcal challenge was performed as previously described (15, 28). Baseline nasal wash and serum samples were taken up to seven days before inoculation. Inoculation entailed installation of an estimated 80,000 CFU per nostril of *Streptococcus pneumoniae* serotype 6B (strain BHN418, GenBank accession number ASHP00000000.1) using a micropipette, with the participant in a reclining chair. Participants recorded and communicated their temperature to the research team by SMS every day for the following week. Nasal washes were repeated on days 2, 7, 9, 14, 22 and 29 post-inoculation, with a second serum sample on day 29. After completion, if participants’ nasal wash remained culture-positive at day 22 or day 29, they were treated with oral amoxicillin for three days. Those who had been colonised at any time point were invited to return up to one year later for rechallenge using the same pneumococcal strain, dose and procedure. Repeat nasal washes for the re-challenge study were taken at baseline and 2, 7 and 14 days post-inoculation. The timeline is summarised in FIGURE 1.

**Figure 1:**
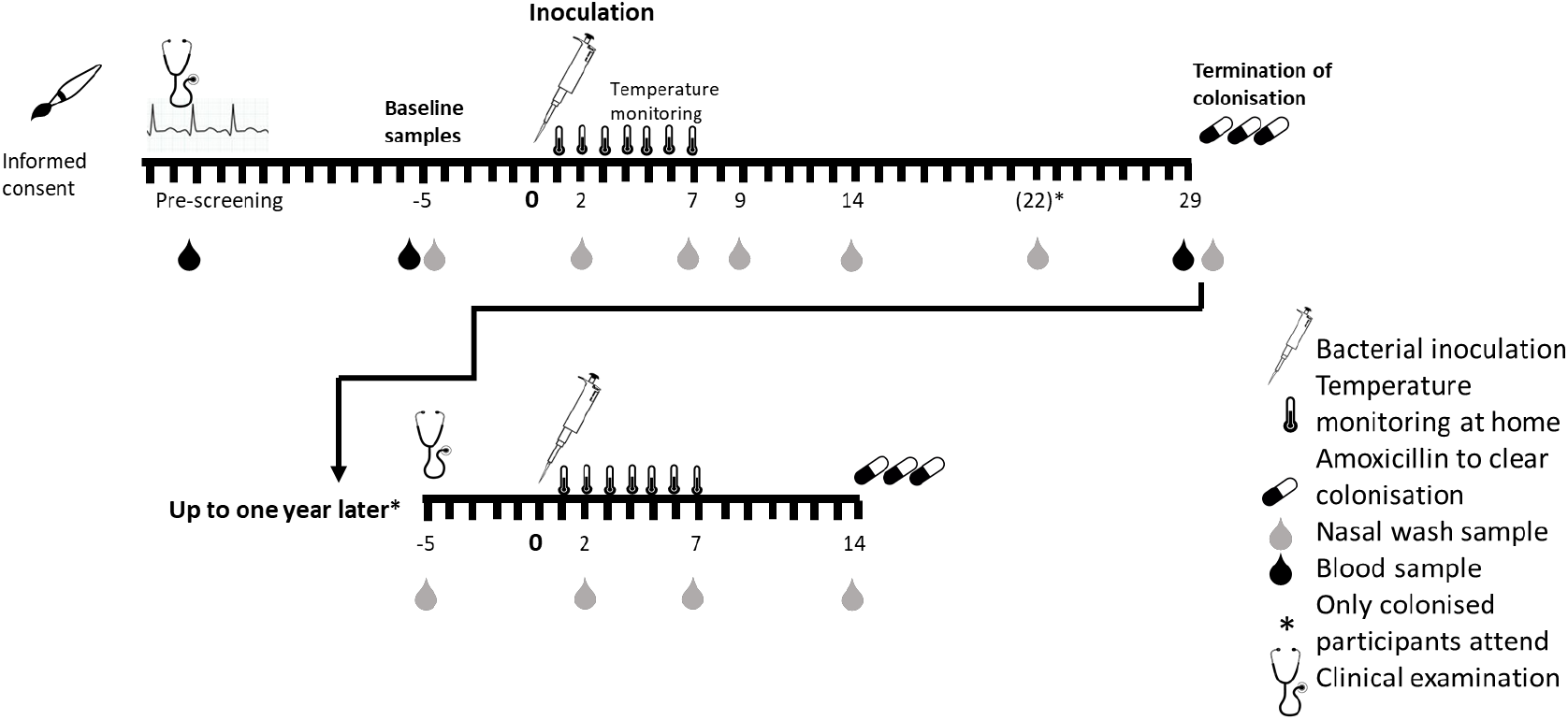
Timeline for the study, including the optional re-challenge (for participants who developed colonisation during the primary study) up to one year later.

Nasal wash samples were processed within an hour of collection, and incubated overnight on gentamicin/blood agar at 37°C in 5% carbon dioxide. Experimental colonisation was defined as the growth of serogroup 6 pneumococcus at any timepoint during the 29 days post-inoculation, identified using standard microbiological criteria (29). Anti-6B capsular polysaccharide (CPS) IgG levels in serum were measured using a modified WHO enzyme-linked immunosorbent assay (ELISA) protocol (see Supplementary Appendix). Serum antibodies against 27 pneumococcal proteins were measured using multiplex electrochemiluminescence (Meso-Scale Discovery, MSD) as previously reported ((15), methodological details in Supplementary Appendix).

### Statistical analysis

The primary endpoint was the rate of experimental colonisation in older adults, which we compared with the rate in younger adults in other EHPC studies using the same methodology carried out during the same time period. We did not recruit a designated young control cohort for this study; >200 young adults took part in other EHPC studies during this time, following the same inoculation protocol.

The typical experimental colonisation rate in young healthy adults is 45% (15) and we hypothesised based on cross-sectional colonisation studies (6) that this would fall to 10% in older adults. A sample size of 64 would detect this difference in rates of experimental colonisation at α = 0.05 with a power of 0.80, allowing for a 10% drop-out rate. We did not pre-specify that we would exclude participants who were naturally colonised with pneumococcus at baseline, but performed post-hoc sensitivity analyses excluding such participants from colonisation outcomes.

Secondary microbiological endpoints included colonisation rates stratified by age, colonisation rates in PPV23-vaccinated participants, colonisation density and duration, and adverse events. Immunological endpoints included the association of pre-existing antibodies with probability of colonisation and colonisation density, and the change in antibody titre after challenge. Total bacterial density during the study was defined as the area under the time-density curve (AUC), calculated according to the trapezoid rule using values of [log_10_(bacterial density+1)] for each interval, with all participants assigned a density of 0 CFU/mL on inoculation day. For participants in the rechallenge phase of the study, colonisation densities up to day 14 post-rechallenge were compared with the colonisation densities over the same time period during the primary challenge. Colonisation rates in different groups were compared using χ^2^ or Fisher’s exact test. Antibody results were log-transformed and compared between groups using the unpaired *t*-test or ANOVA, or within groups (before and after pneumococcal challenge) using the paired *t-*test, with results presented as geometric means and 95% confidence intervals. Correlations between continuous variables were assessed using Pearson’s correlation. Non-parametric tests were used for comparisons of untransformed values within the (smaller) rechallenge cohort. The effects of baseline anti-6B antibodies, adjusted for age and sex, on the development of colonisation or on colonisation density AUC were assessed using logistic regression and linear regression, respectively. Fold changes in the 27 anti-protein antibodies between baseline and day 29 were compared between colonised and non-colonised participants using multivariate regression. No imputations were made for missing data. All tests were two-tailed, and a p value <0.05 was considered significant. All analyses were performed using SPSS version 24 (IBM, New York).

## Results

### Participant characteristics

The oldest participant was aged 80 years, and the median age was 64; the baseline characteristics of the cohort are given in TABLE 1. The recruitment process and screening outcomes are outlined in FIGURE E1 in the Supplementary Appendix. Ten participants were excluded on the basis of abnormal clinical findings at their pre-screening visit, and no participants had previously received pneumococcal conjugate vaccine. Three were naturally colonised with *Streptococcus pneumoniae* at baseline, all of whom remained in the study and underwent experimental inoculation as per protocol.

**Table 1:** Baseline characteristics of all volunteers in the study

|  | Age-defined risk category |  |  | Total |
| --- | --- | --- | --- | --- |
|  | 50-64 | 65-74 | 75 – 84 |  |
| Number of participants inoculated | 34 | 25 | 5 | 64 |
| Number of females | 18 (52.9%) | 15 (60%) | 3 (60%) | 36 (56.3%) |
| Age at inoculation, median (range) | 59 (52—62) | 69 (66—70) | 78 (78—79) | 64 (59—69) |
| Inoculation dose, CFU/mL, median (range) | 83,833 (76,000—90,167) | 84,333 (69,333—92,833) | 84,750 (73,833—89,167) | 84,333 (69,333—92,833) |
| Number of ex-smokers | 10 (29.4%) | 11 (44%) | 2 (40%) | 22 (34.4%) |
| Pack years smoked, median (IQR) | 10 (4.5—10) | 5 (2—10) | 8 (1—8) | 6.5 (3.3—10) |
| Number with any reported comorbidity* | 14 (41.2%) | 15 (60%) | 3 (60%) | 32 (50.0%) |
| Number prescribed any regular medication | 11 (32.4%) | 18 (72%) | 3 (60%) | 32 (50.0%) |
| Prior pneumococcal polysaccharide vaccine | 0 | 17 (68%) | 5 (100%) | 22 (34.4%) |
| Naturally colonised at baseline, n (serogroups) | 3 (23, 3, 15) | 0 | 0 | 3 |

\*Comorbidities reported in more than one participant included the following:
- Benign prostatic hyperplasia: 5 - Depression: 5 - Hiatus hernia: 4 - Hypothyroidism: 4 - Osteoporosis: 3 - Bicuspid aortic valve: 2 - Glaucoma: 2 - Migraines: 2 - Previous malignancy: 2 (1 melanoma, 1 testicular)

### Experimental colonisation rates

The median inoculation dose was 84,333 CFU/mL (range 69,333—92,833). Experimental colonisation developed in 25 participants (39.1%; 95% CI 28.1—51.3%) (FIGURES 2 and 3, and TABLE E3 in the supplementary appendix). This was not significantly different from the 46.7% achieved in 225 young adults in the comparison group (p = 0.281) (FIGURE 3). Colonisation rates within the over-50 cohort did not differ by age decile (χ^2^ for trend p = 0.146), FIGURE 3. When baseline natural carriers were excluded, the overall colonisation rate was 37.7% (26.6—50.3%, n = 23/61).

**Figure 2:**
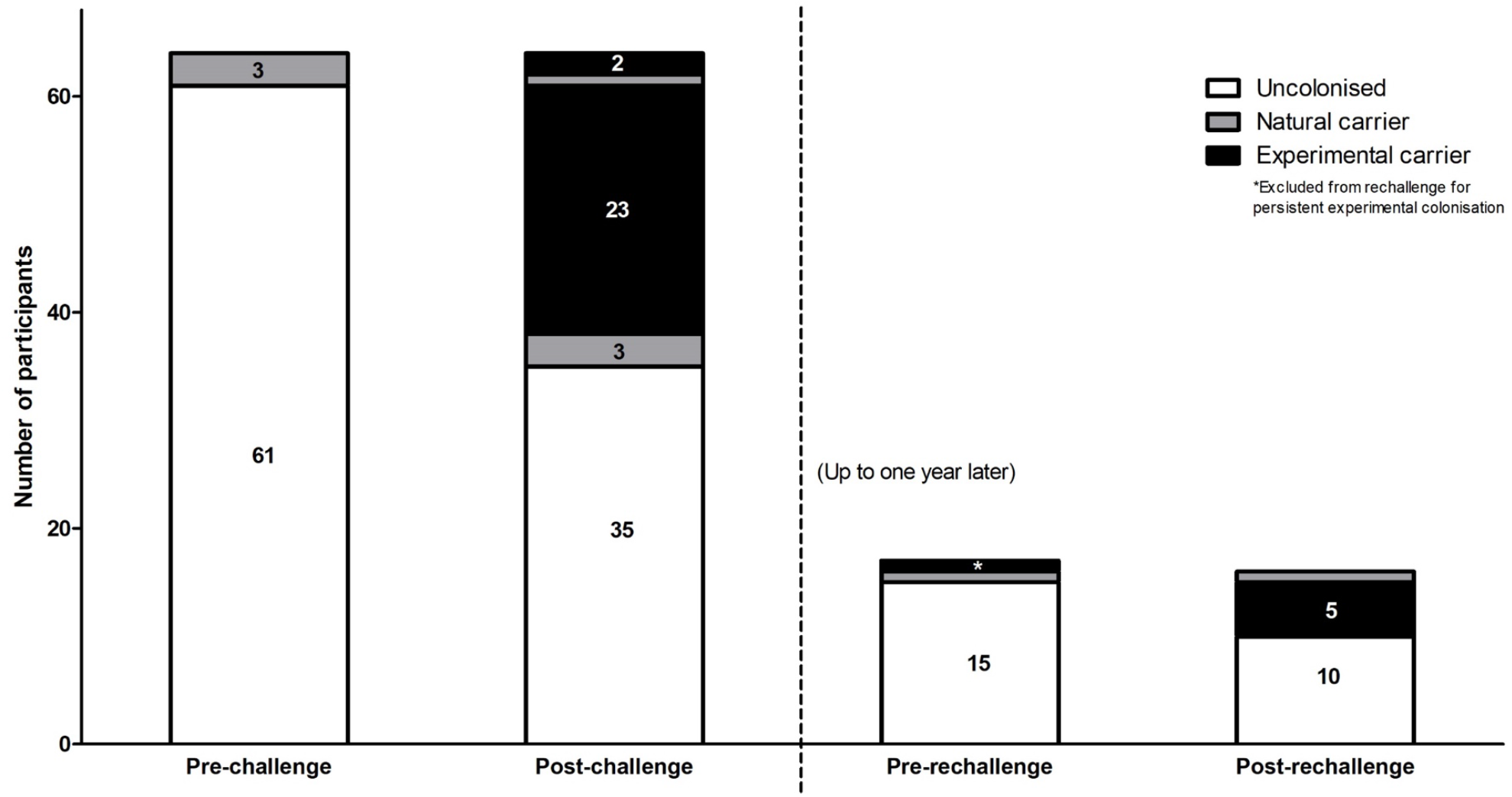
Microbiological status of participants pre- and post-inoculation. Three participants were naturally colonised at baseline, two of whom subsequently developed experimental colonisation following inoculation; 23 previously uncolonised participants developed experimental colonisation, and colonisation with non-experimental strains (serogroups 15, 20 and non-vaccine-type, group G). At screening prior to rechallenge up to one year later, one volunteer was naturally colonised (with serogroup 15) and another was still colonised with the experimental strain; the latter was treated with amoxicillin and excluded from the remainder of the study. Five out of the remaining 16 developed experimental colonisation following rechallenge.

**Figure 0:**
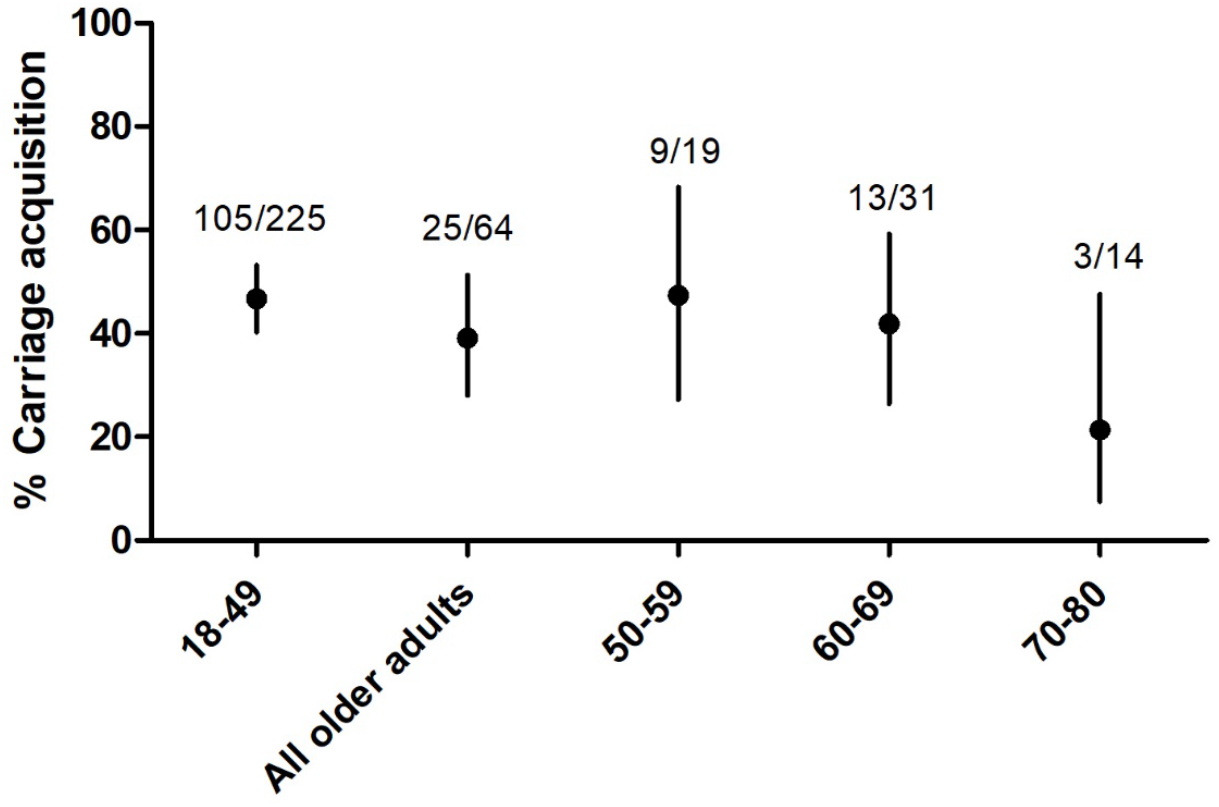
Experimental colonisation rates in older adults (defined by positive pneumococcal culture in nasal wash at any timepoint post-inoculation) compared with a young adult cohort (from similar studies conducted during the same time period), and broken down by age decile within the older cohort. Numbers denote (number colonised)/(total number in that age category). Error bars represent 95% CI. There were no statistically significant differences between the older age category or sub-categories and the younger volunteers.

#### Experimental colonisation density and duration

Colonisation densities varied substantially within and between participants; the average AUC of density was 34.4 CFU.days/mL (95% CI 19.9—48.9) and a graph of individual participants’ colonisation dynamics is in the Supplementary Appendix (FIGURE E2). The median duration of colonisation was 22 days, with 8/25 participants still having detectable colonisation at day 29. Four participants had no detectable colonisation after day 2. AUC of density did not differ with age, either as assessed by age deciles (ANOVA p = 0.84), or by linear correlation with age (r = -0.099, p = 0.64).

#### Experimental colonisation in PPV23 recipients

Experimental colonisation rates were not different in those who were previously immunised with PPV23 compared with those who were not (8/22 [36.4%] vs 17/42 [40.5%] respectively, p= 0.75). This pattern remained in sensitivity analyses restricted to over-65s and to the most recent vaccine recipients. We did not identify any significant predictors of colonisation using logistic models, having tested PPV23 immunisation status and other clinical and laboratory data (Supplementary Appendix).

### Immunological results

Paired serum samples from baseline and 29 days were available for 62/64 participants. There were no differences in baseline antibody anti-CPS IgG titres between males and females (geometric mean 2.82 μg/mL [95% CI 1.91—4.18] and 2.71 μg/mL [1.99—3.69] respectively, p = 0.87). Previous PPV23 was associated with higher baseline antibody levels compared with no prior PPV23 (geometric mean 4.17 μg/mL [95% CI 2.84—6.12] versus 2.24 μg/mL [1.67—2.99] respectively, p = 0.01). We did not find evidence of a fall in antibody levels over time following vaccination. In fact, there was a moderate positive correlation between PPV23 recipients’ baseline antibody levels and time since vaccination (r = 0.46, p = 0.034). Baseline antibody levels increased with age (r = 0.34, p = 0.007), but this correlation was not evident when PPV23 recipients were excluded (r = 0.19, p = 0.24; FIGURE E3 in the Supplementary Appendix).

### Anticapsular antibodies and experimental colonisation

Higher baseline antibody levels were not associated with protection against experimental colonisation: geometric mean IgG concentrations in those colonised and non-colonised were 2.71 μg/mL (1.91—3.84) and 2.79 μg/mL (2.0—3.9) respectively (p = 0.90). Adjustment for age and sex using logistic regression did not alter these findings (Supplementary Appendix).

We did not observe an increase in antibody levels following colonisation (FIGURE 4, TABLE E5). Of interest, pneumococcal challenge without colonisation led to diminished antibody levels: the geometric mean titre in subjects who did not develop experimental colonisation fell from 2.79 μg/mL (2.9—3.9) at baseline to 2.17 μg/mL (1.57—3.0) at day 29 (p = 0.006). On sensitivity analyses subdivided by PPV23 administration, this finding was significant only in the vaccinated sub-cohort (FIGURE 4; TABLES E6 and E7 in the SUPPLEMENTARY APPENDIX).

**Figure 0:**
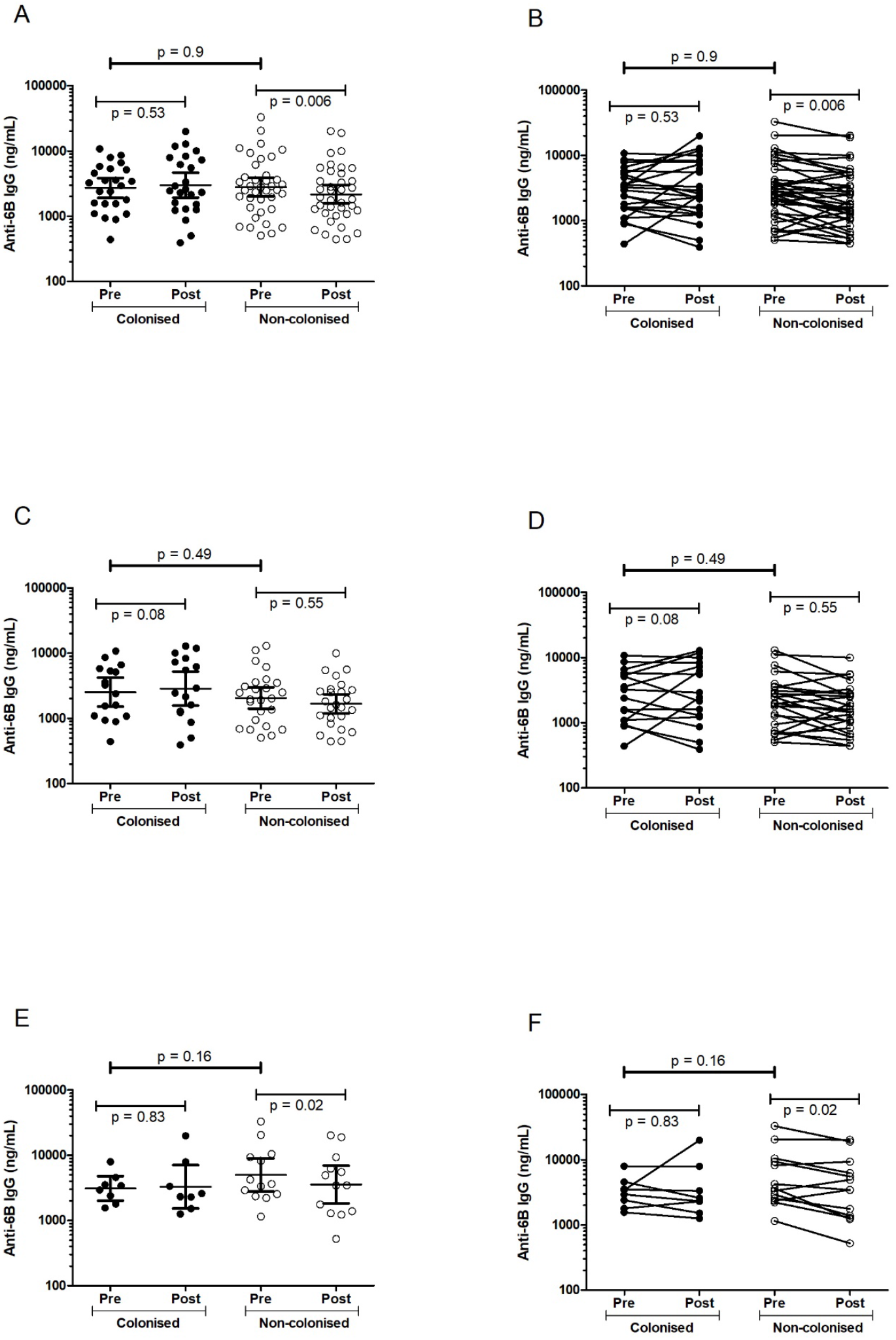
Anti-6B CPS IgG levels at baseline and day 29 following inoculation, in the full cohort (A, B), participants who had never received PPV23 (C, D) and PPV23-vaccinated participants (E, F). Each symbol represents a single participant. The lines and error bars in A, C and E represent geometric mean (95% CI); the lines in B, D and F connect the baseline and day 29 values for each participant.

When antibody responses in colonised and non-colonised participants within each age decile were compared, the only statistically significant change was the fall in antibody levels post-challenge in non-colonised over-70s (all of whom had received PPV23; TABLE E8 in the SUPPLEMENTARY APPENDIX). However, the statistical significance was borderline (p = 0.047) and we did not correct for multiple comparisons. When antibody responses to colonisation were summarised as fold changes, there was no correlation with age in either colonised (r = -0.025, p = 0.906) or non-colonised participants (r = -0.155, p = 0.352). Higher baseline antibody levels did not correlate with lower colonisation density, nor did higher density correlate with higher day-29 antibody levels or greater fold change in antibody levels (SUPPLEMENTARY APPENDIX).

### Anti-protein antibodies

Serum antibody levels against 27 pneumococcal proteins were measured at baseline and at day 29 post-challenge using MSD multiplex electrochemiluminescence. There were no differences in the baseline levels of any antibody between colonised and non-colonised participants (FIGURE E4). In contrast to the anti-capsular IgG response, antibody titres against several pneumococcal proteins were increased following pneumococcal colonisation, including PspC, PspA-UAB055, RgrA-Tigr4, PiuA and PcpA (all p<0.001) (FIGURE 5). In individuals who remained uncolonised, there was no significant change in any anti-protein antibodies.

**Figure 5:**
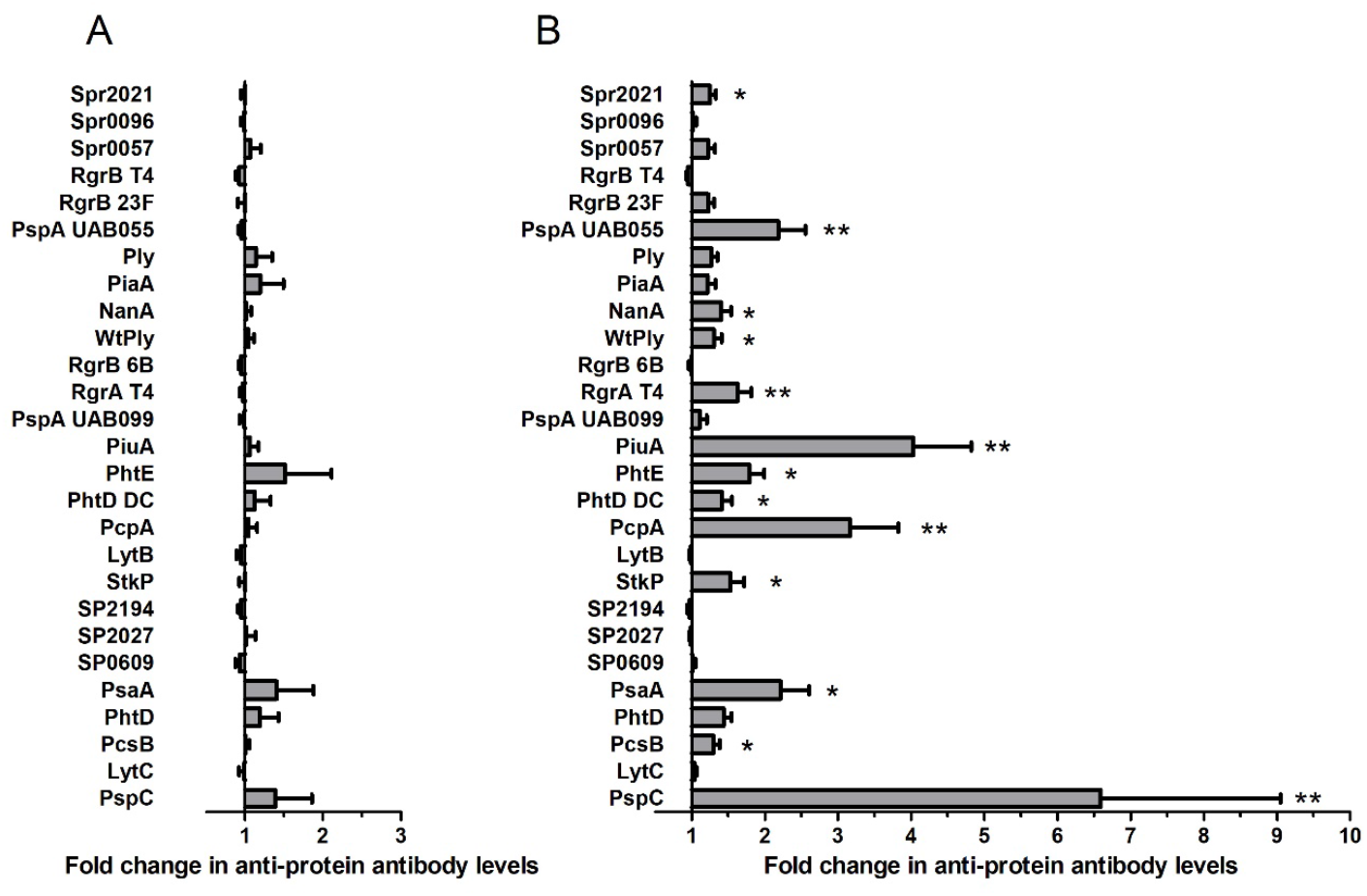
Changes in antibody levels against 27 different pneumococcal proteins following nasopharyngeal pneumococcal challenge, measured by multiplex electrochemiluminescence (Meso-Scale Discovery), in non-colonised (A) and colonised (B) participants. Antibodies are expressed as fold change between pre-challenge baseline and 29 days post-inoculation with S pneumoniae. Error bars represent mean (SEM). Significant differences in fold change between carriers and non-carriers were analysed using multivariate regression: * denotes p < 0.05, ** p < 0.001.

### Mucosal antibodies

Anti-6B antibody levels in nasal wash were similar at baseline between colonised and non-colonised participants, and these levels did not change significantly following bacterial challenge (FIGURE E5)

### Protection against experimental re-colonisation

Rechallenge was performed in 16 of the 25 participants who originally developed experimental colonisation, after a median interval of 259 days (IQR 205—308 days; Figure S1 in the Supplementary Appendix). One was colonised with serogroup 15 pneumococcus at time of rechallenge, and participated in the study as per protocol. A further participant was colonised with a serotype 6B pneumococcus at time of rechallenge; he could not recall whether he had taken the 3- day course of amoxicillin prescribed following the initial study completion. Community-acquired colonisation with serogroup 6 pneumococci is uncommon in Liverpool (30). The participant was not re-inoculated, but treated with amoxicillin for five days. Subsequent nasal washes at seven and 28 days were negative by both culture and polymerase chain reaction.

The median age of rechallenge participants was 63.5 years, and 11 (68.7%) were female. Within 14 days of inoculation, colonisation was detected in five participants (31.3%).

### Colonisation density in rechallenge participants

Of the five colonised participants, two had detectable colonisation at all three post-inoculation timepoints, while the remainder were only culture-positive at one timepoint each. The mean colonisation density by day 14 in rechallenge participants (as measured by AUC) was 10.0 CFU.days/mL (95% CI 0—21.94). This was non-significantly lower than the mean density in the same five participants during the first 14 days of the primary challenge (31.01 CFU.days/mL, 95% CI 11.08—50.95, p = 0.08 using Wilcoxon’s matched pairs test).

### Anti-capsular antibodies in rechallenge participants

Antibody levels pre- and 14 days post-rechallenge are shown in FIGURE 6, and in TABLE E11 in the SUPPLEMENTARY APPENDIX, along with the corresponding antibody levels from the same participants in the primary challenge for comparison. One participant did not have serum taken on day 14 post -rechallenge. Antibody levels did not fall significantly during the interval between the end of the primary challenge and the start of the rechallenge. The pre-rechallenge baseline antibody levels were similar between participants who did and did not develop colonisation following rechallenge: respective GMTs (95% CI) 3.72 μg/mL 1.18—11.76) and 1.66 μg/mL (1.17—2.36) (p = 0.07 using the Mann-Whitney U test). Antibody levels fell in participants who remained uncolonised post-rechallenge (GMT at day 14 1.53 μg/mL (95% CI 1.08—2.17), p = 0.041 using Wilcoxon’s signed rank test to compare with pre-challenge levels).

**Figure 6:**
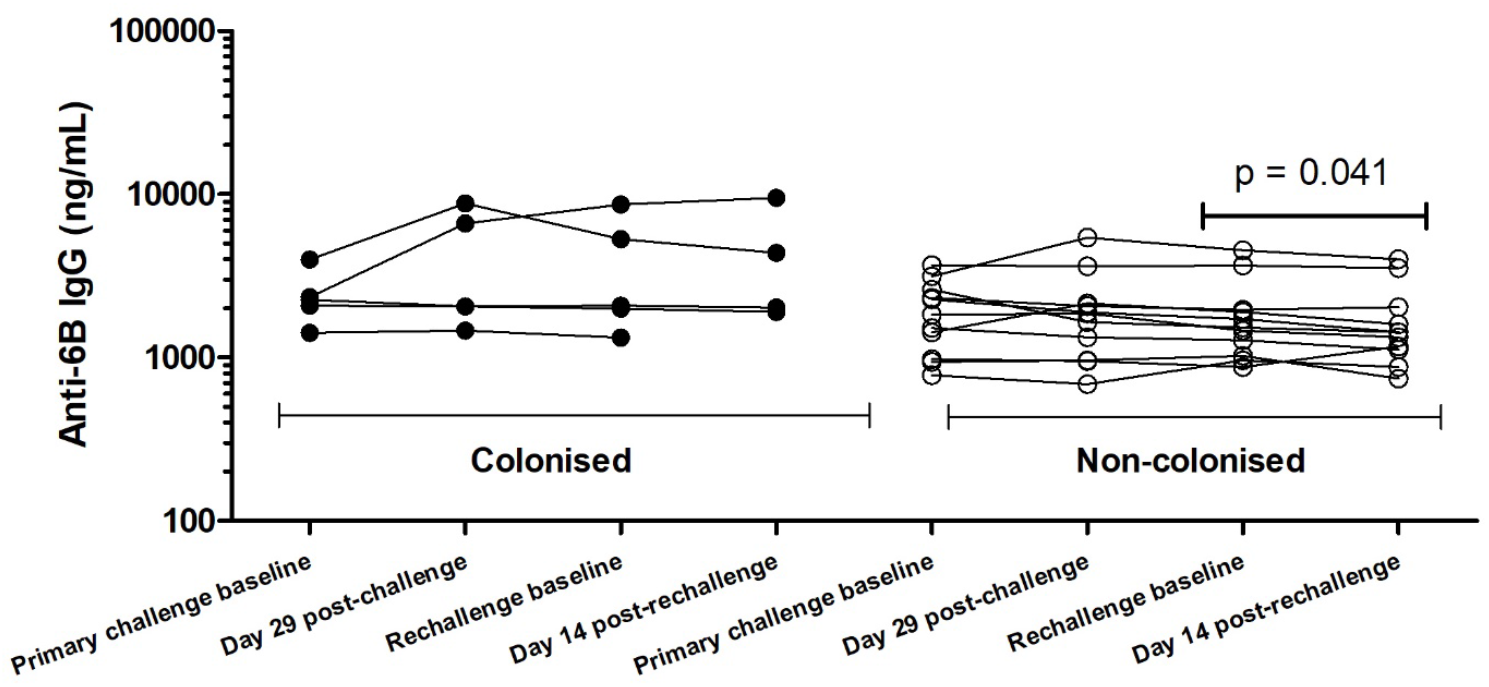
Serum anti-6B capsular antibodies before and after the primary and rechallenge phases of the study. Only participants who were colonised following the primary challenge and returned for rechallenge are shown, subdivided into those who did and didn’t become colonised following rechallenge. Each dot represents an individual participant.

### Safety of EHPC in older adults

There were no serious adverse events, and no cases of pneumococcal disease. During active surveillance (the week following inoculation), no participants recorded oral temperatures ≥38°C. During the primary challenge, seven participants developed symptoms which, when reviewed clinically, were deemed unrelated to study procedures by the investigators; details are summarised in the Supplementary Appendix. During the rechallenge phase, one colonised participant developed malaise and unilateral otalgia at day 11, but sought no medical help. They attended routinely at day 14, at which time clinical examination and otoscopy were unremarkable. For precaution, a five-day course of amoxicillin was prescribed to clear colonisation.

## Discussion

This is the first human bacterial challenge model in older adults. Pneumococcal colonisation was safely established at similar rates, densities and durations as in younger adults (15, 29). Colonisation rates in the oldest participants in this cohort (ages 70—80 years) were non-significantly lower than those seen in under-50s, but low numbers (n=14) preclude definitive conclusions. The ranges and fluctuations in density and variable durations of colonisation were similar to those seen in young participants following both experimental and natural colonisation (29, 31). PPV23 receipt did not protect against colonisation, and baseline antibody levels were not different in colonised and non-colonised participants, the latter observation consistent with previous findings in young adults (15).

However, immunological outcomes following pneumococcal challenge differ considerably from young adults, whose anticapsular antibody levels are raised following experimental colonisation remain and unchanged in non-colonised participants (15). Amongst older adults, antibody levels were unchanged following colonisation and actually fell in non-colonised participants. Our statistical power to detect age-related immunological trends within the cohort itself was limited by small numbers in each age decile. During rechallenge we observed no protection against re-acquisition of homologous serotype colonisation in this older cohort in contrast to the universal protection seen in a previous study of participants aged <50 years (0/10 recolonised) (15).

There are a number of potential explanations for the antibody responses we describe in older adults. Serotype-specific memory B cell levels have been shown to fall following revaccination with PPV23 in older adults (32), and memory B-cells may undergo similar terminal differentiation in response to live pneumococcus. Our results may reflect those of vaccine studies which have demonstrated hyporesponsiveness following repeated doses of polysaccharide in the form of PPV23 (33-35). This hypothesis is supported by our finding that antibody levels fell more in PPV23-vaccinated participants than in unvaccinated participants following challenge, although numbers were small in these subgroups, and we did not correct for multiple comparisons. The hyporesponsiveness hypothesis is further supported by the fall in anti-CPS antibodies in non-colonised participants following rechallenge. Alternatively, peripheral antibodies were perhaps sequestered in the nose following pneumococcal challenge, leading to a drop in circulating levels. We have previously reported mucosal antibody sequestration by inoculated bacteria in PCV13-vaccinated young adults (preventing colonisation via agglutination), but did not observe an effect on peripheral antibody levels (24). Equally, in the current study we did not identify any changes in nasal wash anti-6B IgG levels at any timepoint post-challenge in colonised or non-colonised participants.

Our finding that PPV23 does not prevent colonisation contrasts with a recent study in US adults aged ≥65 years. The authors followed 100 participants with fortnightly sampling for one year and identified a high period prevalence of colonisation in the cohort as well as significantly lower colonisation in participants who had ever received either pneumococcal vaccine (36). However, most pneumococcal detections were by *lytA* qPCR rather than culture, and serotype confirmation was unavailable for the vast majority of isolates. Longitudinal surveillance of US children suggests that any reduction in vaccine-type pneumococcal colonisation following vaccination is mostly balanced out by serotype replacement (37). It seems unlikely that PPV23 vaccination of older adults disrupts colonisation to such a degree.

Using MSD, we identified boosted antibody levels against a number of pneumococcal proteins following colonisation. These included PspA-UAB055, PspC and PiuA, which were also boosted following colonisation in young adult participants in a previous study, using the same MSD methodology (15). This suggests that these proteins are visible to the immune system during colonisation, and are good markers of exposure. Our finding that baseline anti-protein antibody levels were not different between colonised and non-colonised participants suggests that serum anti-protein immunity may not be a major component of anti-pneumococcal immunity in older adults.

### Limitations

Our participants were carefully screened, which may have resulted in a particularly healthy cohort, characterised by non-smoking and minimal medical comorbidity. While this limits the generalisability of our findings, we felt it was necessary to maximise the safety of participants. We defined colonisation based on culture positive nasal wash samples, rather than molecular testing with typically higher sensitivity (31, 38, 39). Some authors have argued that the oropharyngeal niche has higher yield for detecting colonisation in older adults than our nasal wash method (9). We did not measure functional antibody activity in this study, but IgG levels and serum opsonophagocytic activity are reasonably correlated in older adults (19). The antibody response to colonisation may be serotype dependent: population studies have shown a decline in antibodies against certain serotypes (e.g. serotype 3) with age, but not in other serotypes including 6B (40). By contrast, an observational study in young adults found that 6B colonisation did not elicit as strong an antibody response as other serotypes (14). Future research should study the immune response of older adults to challenge with non-6B serotypes.

## Conclusions

New strategies are needed to protect older people against pneumococcal disease. The safety, tolerability and high rates of experimental colonisation seen in this study support the use of EHPC for vaccine testing in this key vulnerable population.

## Data Availability

N/A

## Acknowledgements

We thank all study participants for their time and commitment. In addition, we thank Catherine Molloy, Kelly Convey, John Blakey, Hassan Burhan, Ben Morton, the members of the Data Safety and Monitoring Board (Robert Read and Brian Faragher), the Clinical Research Network North West Coast, the governance staff of the Liverpool School of Tropical Medicine and the Royal Liverpool and Broadgreen University Hospitals NHS Trust and the Royal Liverpool University Hospital Clinical Research Unit.

